# Undermining Fatigue in Chronic Conditions

**DOI:** 10.1101/2025.07.01.25330359

**Authors:** Rosalie M. Grijalva, Curtis J. Perry, Rachel J. Perry

## Abstract

While progress has been made toward understanding physical and mental fatigue, chronic fatigue has been under-studied and stigmatized. Unlike other fatigue states that can be relieved by rest, chronic fatigue is a common, debilitating symptom of many chronic conditions. Utilizing deidentified patient data from the Yale-New Haven Hospital System, we analyzed the overlap between diagnosed fatigue and 14 chronic conditions. Our results revealed a significantly lower overlap with fatigue than previous reports in all but one of the current diagnoses’ fields. The underreporting of disabling fatigue across chronic conditions restricts the translation between medical and basic scientific research. Accurate reporting of the prevalence of chronic fatigue can result in researchers refocusing their efforts toward uncovering targetable mechanisms and physicians consistently reporting and treating chronic fatigue.

---

One of the first investigations into the mammalian immune system described a strikingly similar physiological response to a diverse range of harmful stimuli in rats.^1^ An unfamiliar biological stimulus, such as an infection, consistently triggered specific animal reactions, including anorexia, fever, depression, and fatigue, which were later termed “sickness behaviors.”^1,2^ Sickness behaviors reallocate energy to critical processes like inflammation and thermoregulation.^2^ Sickness behaviors can be beneficial following acute insults, yet the same responses are detected and can be disabling in chronic illness.

The importance of elucidating sickness behavior mechanisms and effective treatments is heightened with the increased prevalence of chronic conditions and substantial associated health care costs.^4^ While chronic conditions may present differently, they share ubiquitous sickness symptoms akin to animal sickness behaviors. Across chronic obstructive pulmonary diseases, chronic heart failure and end-stage renal disease, a majority of patients reported three persistent symptoms: pain; numbness; and a lack of energy.^5^ While each of these symptoms illustrates a critical need, “lack of energy” or chronic fatigue, is a debilitating symptom identified across chronic conditions with no effective treatments.

Chronic fatigue, and its connection to other conditions, is underappreciated in scientific and medical fields and represents an important area for future research to improve the quality of care for those with chronic illness. One explanation for the lack of research on chronic fatigue is the systemic underreporting of chronic fatigue, muting the urgency of investigating this quality of life-altering condition. To investigate the local prevalence of diagnosed chronic fatigue across a range of chronic illnesses, we used Slicer/Dicer software to access generalized medical diagnoses among deidentified patients in the Yale-New Haven Hospital (YNHH) system.

We searched the database for the overlap of fatigue and 14 chronic medical diagnoses: multiple sclerosis (MS), traumatic brain injury (TBI), fibromyalgia (FM), Alzheimer’s disease (AD), glioblastoma (GB), myasthenia gravis (MG), amyotrophic lateral sclerosis (ALS), chronic kidney disease (CKD), chronic obstructive pulmonary disease (COPD), obesity (OB), type 1 and type 2 diabetes (D1 and D2, respectively), cystic fibrosis (CF) and metastatic melanoma (ML). These conditions were selected based on the chronic nature of the disease, prevalence, and the association with fatigue. We aimed to characterize fatigue in diverse chronic illnesses from rare diseases, like GB and FM, to more common conditions, such as OB and AD. Of the investigated diseases, MS^6,7^, FM^8^, AD^9,10^, COPD^11^, and OB^12^ are commonly associated with persistent fatigue. Several of the other diseases have a weaker known association with fatigue. For example, exercise, depression, and fatigue have been briefly studied in CF, a disease typically caused by a transporter mutation that affects lung function and digestion.^13,14^

In the YNHH system, the total number of patients identified across all disease search terms was 521,662, with approximately 11% of patients diagnosed with fatigue alongside one of the 14 chronic conditions we investigated (**Figure 1A**). These results were compared to the total number of patients in the YNHH system (*n*=4,756,338) and the patient number with a medical history, diagnosis or chief complaint of fatigue (*n*=114,152). The total rate of reported fatigue is 2% of all patients in the YNHH, which is dramatically lower than the 11% fatigue documented in the conditions we investigated (**Figure 1A; S1**). These results demonstrate that chronic fatigue is more reported in chronic conditions in the YNHH system.

**Figure 1.**
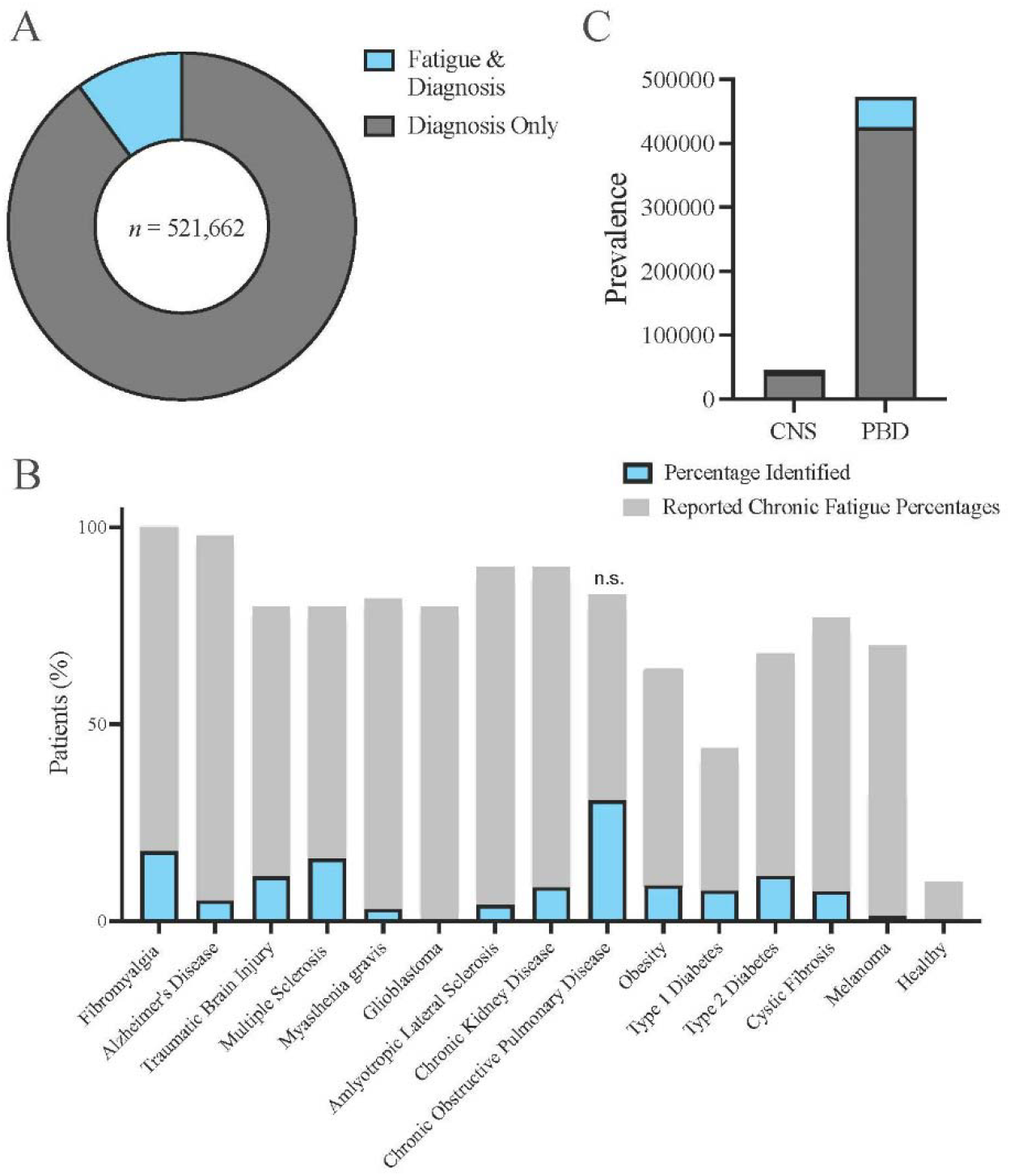
In the Yale New Haven Hospital System, the rate of diagnosed fatigue in chronically ill patients is 11% and dramatically lower than previous reports. Slicer/Dicer software was utilized to assess diagnosed fatigue in 14 prevalent chronic illnesses. A) Total patients under the searched terms were assessed (n=521,662) and revealed an overlap of chronic condition diagnoses and fatigue at 11%. B) Separating the chronic conditions, the resulting overlapping percentages were compared to a range of previous percentages of patients reporting chronic fatigue with the same disorders. Light gray represents the range of previously reported percentages. C) Conditions were separated into two categories based on primary systems impacted:

To further assess fatigue prevalence, we next calculated the individual percentage of overlap between fatigue and another diagnosis over the total patients with the respective chronic condition. The highest overlap with fatigue was 30% in patients with COPD, a trending but insignificant difference to a previously reported frequency of 55% of COPD patients **(n.s; Figure 1B**).^11^ The rest of the chronic diagnoses were markedly decreased compared to previous fatigue reports. Surprisingly, we found a mere 18% overlap with FM and fatigue, a significant decrease compared to several reports have identified a fatigue prevalence in patients with FM at around 77% (p<0.0001; **Figure 1B**).^8,16^ A diagnosis of GB had the lowest fatigue overlap of each of the disorders at significantly less than 1% of YNHH cases. However, previous reports estimate that GB patients have a substantial incidence of fatigue at 48%^17^ and general brain tumors have an overwhelming fatigue rate of approximately 80%^18^ (p<0.0001; **Figure 1B**). For reference, we plotted the previously reported fatigue ranges in each chronic condition alongside our results (**Figure 1B**). Our identified overlap between each respective chronic diseases and fatigue was significantly lower than previous reports in 13 of the 14 current diagnoses’ fields (p<0.0001).

To assess if chronic fatigue is diagnosed more consistently in diseases originating in the central nervous system (CNS) or the peripheral body (PBD), we separated the 14 chronic diseases according to the major system impacted. The CNS category consisted of MS, TBI, FM, AD, and GB. The PBD group included CKD, COPD, OB, D1, D2, and CF. The prevalence of PBD patients greatly outnumbered the CNS patients, with OB and D2 representing the most diagnosed conditions (**Figure 1C**). Despite the prevalence gap, the PBD category averaged 11.2% overlap with fatigue compared to a 11.7% average in CNS diseases, suggesting that fatigue is similarly diagnosed regardless of the system impacted.

Central Nervous System (CNS) and Peripheral Body Diseases (PBD). CNS disease include fibromyalgia (FB), Alzheimer’s disease (AD), traumatic brain injury (TBI), multiple sclerosis (MS), glioblastoma (GB), and amyotrophic lateral sclerosis (ALS). PBD are chronic kidney disease (CKD), chronic obtrusive pulmonary disease (COPD), obesity (OB), type 1 and type 2 diabetes (D1 and D2, respectively), cystic fibrosis (CF) and metastatic melanoma (ML). Across panels, blue illustrates our overlap of fatigue and the respective chronic illness. Dark grey represents the presence of a chronic condition and lack of a fatigue diagnosis. N.S. stands for non-significant. All other comparisons were statistically significant.

While difficult to put an overall expected percentage of chronically ill patients that suffer from fatigue, it is likely higher than our reported 11%. To support this conclusion, a recent meta-analysis identified a global fatigue prevalence in healthy individuals at 11.2%, likely representing individuals with unexplained chronic fatigue or chronic fatigue syndrome (CFS).^15^ Strikingly, our data suggests that patients with chronic illness experience the same rate of documented chronic fatigue as healthy individuals. While we are comparing different populations with varied demographics, it is increasingly likely that many of these chronically ill patients confront fatigue on a daily basis. Furthermore, in the total patient population in the YNHH system the rate of fatigue was 2% (**Supplementary Figure 1**), lower than the 11% global fatigue rate in healthy individuals. Our results suggest a general underreporting of chronic fatigue across the YNHH system.

While the percentage of overlap between the diverse conditions and fatigue ranged from less than 1% to 30%, diagnosed fatigue for 13 of the chronic illness were significantly decreased compared to previous accounts, exhibiting a widespread under-reporting of fatigue in chronic conditions in the YNHH system (*p*<0.0001). One rationale for the discrepancy between our results and the literature is the lack of fatigue diagnosis, even when symptoms are present. This may be due to a de-prioritization of fatigue as compared to life-threatening diseases, or a focus towards symptoms with known or more effective treatments. Additionally, if fatigue is a known symptom of a chronic condition, a diagnosis of fatigue may appear redundant. However, in the translation between medical and basic scientific fields, the lack of fatigue diagnoses in chronically ill patients limits the understanding of fatigue across conditions and decreases the chances of fatigue treatment for these patients.

While there is no known mechanism behind chronic fatigue across conditions, there is evidence of immune system dysregulation, metabolic alterations, and organismal changes in the brain and body of patients suffering from CFS.^19^ Dissecting this mechanism further requires promising animal models, precise methods, and investigation into brain-body interactions in fatigue. Healthy metabolism requires proper interaction between the brain and the body to sense and respond to energetic supply and demand. Although the mechanisms underlying fatigue are not fully understood, it is well established that the best current treatment for fatigue is exercise. Exercise is generally considered to be a simple manipulation, but for chronic fatigue patients it can be extremely difficult to “find the energy.” Therefore, rather than pushing chronically ill patients to exercise, future research is critical to elucidate fatigue mechanisms. These mechanisms may apply to fatigue in many diverse contexts to better our understanding of general health and disease. Elucidating metabolic mechanisms across the brain and body may result in researchers refocusing their efforts toward uncovering targetable mechanisms and physicians consistently reporting and treating chronic fatigue and its metabolic underpinnings.

## Methods

Slicer/Dicer software was used to access generalized medical diagnoses among deidentified patients in the Yale-New Haven Hospital (YNHH) system. IRB approval is not needed for Slicer/Dicer data. The database was searched using the following terms: “multiple sclerosis,” “traumatic brain injury,” “fibromyalgia,” “Alzheimer’s disease,” “glioblastoma,” “myasthenia gravis,” “amyotrophic lateral sclerosis,” “chronic kidney disease,” “chronic obstructive pulmonary disease,” “obesity,” “type 1 diabetes,” “type 2 diabetes,” “cystic fibrosis,” and “metastatic melanoma,” and “fatigue.” The database was only able to retrieve diagnoses, and did not have access to patient notes. These conditions were selected based on the chronic nature. Each diagnosis percentage was compared to the lowest reported percentage in the respective condition.

## Statistics

A two tailed binomial test with patient percentages was utilized in Prism 10.5 to assess significance. Wilson/Brown test determined the confidence interval. The expected input to be compared to our results consisted of the lowest patient percentage of fatigue in previous reports. n.s. nonsignificant. \*\*\**p*<0.0001.

## Data Availability

All data produced in the present work are contained in the manuscript.

## Extended Data

**Supplementary Figure 1:**
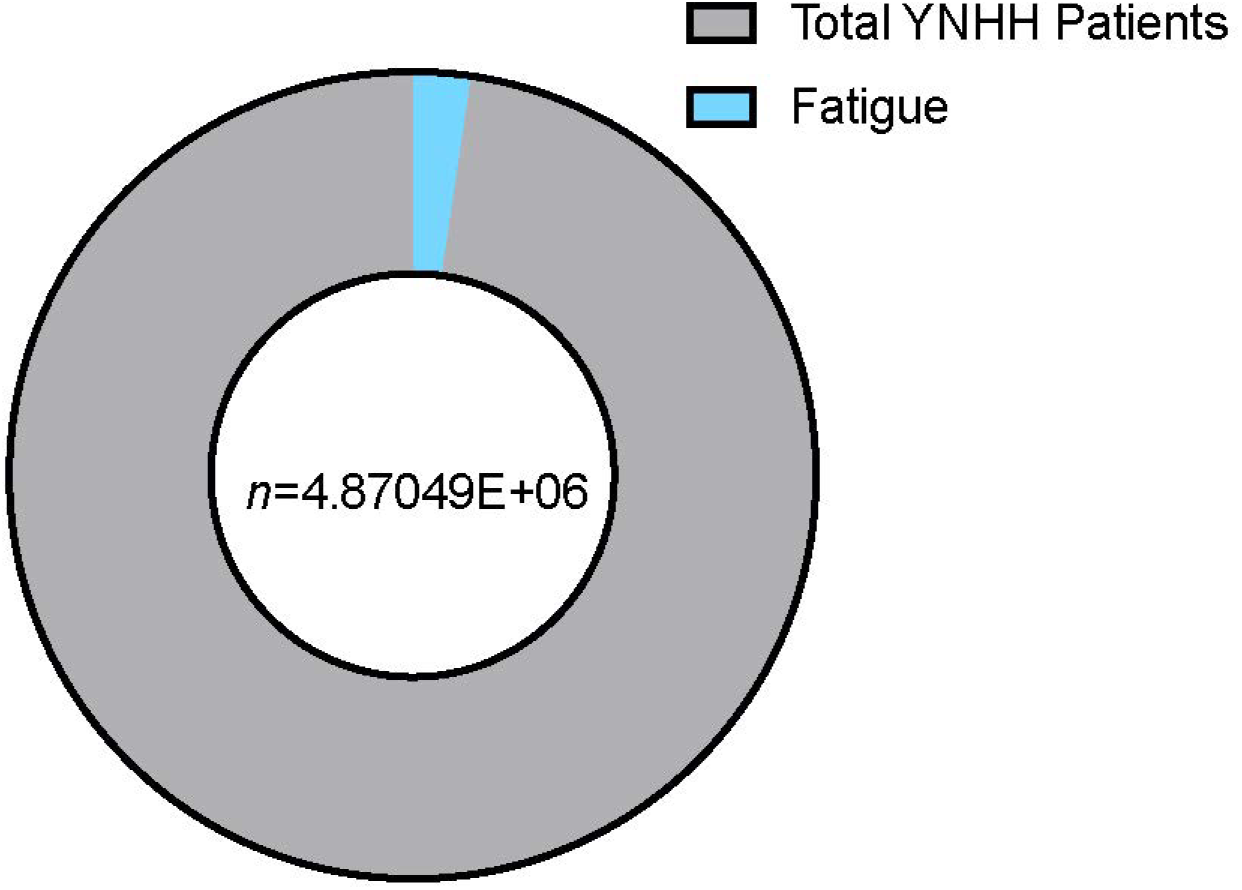
In the complete patient population in the Yale New Haven Hospital (YNHH) System only 2% are reported to suffer from chronic fatigue. Slicer/Dicer software was utilized for all of the YNHH patients (*n*=4,756,330, light grey). The light blue represents reported fatigue in the complete population.

